# *Plasmodium vivax* and *Plasmodium falciparum* mixed infections in human and mosquito hosts: the impact of multi-species infection on parasite densities and transmission to mosquitoes

**DOI:** 10.64898/2026.03.16.26348472

**Authors:** Wakweya Chali, Legesse Alamirie Ejigu, Tigist Atele, Solomon Sisay, Gutema Jebessa, Getnet Habtamu, Melat Abdo, Amanuel Shimelash, Mulugeta Demisse, Migbaru Keffale Bezabih, Desalegn Nibret, Addisu Gizat, Kebede Getachew, Kassahun Abi, Ayalew Jejaw Zeleke, Banchayehu Getnet, Zewdu Solomon, Teressa Bayena, Tirhas Endale, Meskele Dechasa, Fekadu Massebo, Fikregabrail Aberra Kassa, Sagni Challi, Hassen Mamo, Chris Drakeley, Teun Bousema, Fitsum G. Tadesse

**Affiliations:** Armauer Hansen Research Institute, Addis Ababa, Ethiopia; Radboud University Medical Centre, Nijmegen, The Netherlands; London School of Hygiene and Tropical Medicine, London, United Kingdom; Department of Biology, Arba Minch University, Arba Minch, Ethiopia; Department of Microbial Sciences and Genetics, College of Natural and Computational Sciences, Addis Ababa University, Ethiopia; Department of Medical Parasitology, School of Biomedical and Laboratory Sciences, University of Gondar, Gondar, Ethiopia

**Keywords:** mixed infections, gametocytemia, transmission, membrane feeding assay, *P. vivax*, *P. falciparum*, *Anopheles*

## Abstract

In co-endemic regions, mixed *Plasmodium vivax (Pv)* and *Plasmodium falciparum (Pf)* infections are commonly reported. How mixed species infections compare to single species infections in terms of parasite densities and transmission to mosquitoes is incompletely understood. Parasitemia, gametocytemia, and mosquito infectivity were evaluated among *Pv* mono-infections (n=284), *Pf* mono-infections (n=150), and mixed-*Pv-Pf* infections (n=77) recruited at four Ethiopian health facilities. Parasitemia and gametocytemia were quantified in patient blood samples by qPCR. Mosquito infectivity was assessed using direct membrane feeding assays (DMFA), with *Plasmodium* species confirmation by PCR. *Pf* gametocyte prevalence was lower in mixed-infections (64.5%, 49/76) compared with mono-infections (91.8%, 135/147); among gametocyte-positive parasite carriers gametocyte density was also lower in mixed-infections (*p*<0.001). *Pv* gametocyte prevalence was similar in mono- and mixed-infections despite lower asexual parasite density in mixed infections (*p*<0.001). Statistically significant positive correlations between asexual parasitemia and gametocytemia were observed in mono-infections (*p*<0.001), but not in mixed-species infections (*p*=0.120 for *Pv*; and *p*=0.570 for *Pf*). Transmission to mosquitoes was high across infections and infection combinations. Among infectious *Pv-Pf* parasite carriers, 56.3% (27/48) transmitted both species, often with both species being transmitted to individual mosquitoes. The associations between (species specific) gametocyte density and mosquito infection rates was unaffected by the concurrent presence of the other *Plasmodium* species. Although mixed-species infections may have different parasite and gametocyte densities compared to mono-infections, we observed no evidence for competition between species in mosquitoes. Because mixed infections are often undetected, they represent a hidden risk for sustaining malaria transmission.

**Author Summary:** Malaria remains a major public health challenge in Ethiopia, where *Plasmodium vivax* and *Plasmodium falciparum* often occur together in the same patient. While mixed-species infections are common, little is known about how they compare to single-species infections in terms of parasite levels and transmission to mosquitoes. In this study, we examined patients with *P. vivax*, *P. falciparum*, or mixed infections across four health facilities. We measured parasite and gametocyte densities in blood samples and tested whether patients could infect mosquitoes. We found that *P. falciparum* gametocyte levels were lower in mixed infections compared to single-species infections, while *P. vivax* gametocyte levels were similar regardless of infection type. Importantly, patients with mixed infections were still highly infectious to mosquitoes, and many transmitted both parasite species simultaneously. These findings show that mixed infections, which are often missed by routine diagnosis, can contribute substantially to malaria transmission. Recognizing and addressing mixed-species infections is therefore critical for malaria control and elimination efforts.

## Introduction

Malaria transmission in regions where *Plasmodium falciparum* and *Plasmodium vivax* are sympatric is characterized by a striking epidemiological paradox. *P. vivax* incidence and prevalence are often lower than *P. falciparum* [1], despite *P. vivax* possessing several biological traits that theoretically favor persistence and spread. These biological traits include rapid gametocyte production within 2–3 days of blood-stage infection [2], efficient transmission at lower parasite densities [3], relapse-driven replenishment of the infectious reservoir [4], and accelerated sporogonic development across a wide range of temperatures that enables it complete sporogony within the lifespan of the vector. Since *P. vivax* possesses a dormant liver stage (hypnozoites) and individuals are often exposed to both *P. falciparum* and *P. vivax*, treatment of *P. falciparum* infections is frequently followed by subsequent *P. vivax* episodes.

As control interventions increasingly suppress *P. falciparum* transmission, *P. vivax* has emerged as highly important cause of malaria in many endemic regions [5]. Because both species are typically transmitted by the same *Anopheles* mosquitoes, understanding why *P. vivax* does not dominate under favourable conditions remains central to unravelling their shared epidemiology. One potential explanation for the above-described paradox may lie in interactions between co-circulating species. The occurrence of mixed-species infections is increasingly recognized, yet their epidemiological and biological consequences remain poorly understood. Routine surveillance substantially underestimates co-infections due to diagnostic limitations: microscopy detects only 5-20% of mixed *P. falciparum–P. vivax* infections, whereas molecular diagnostics reveal prevalences of up to 30% [6–12].

Contrasting with regular detection of co-infections in human parasite carriers, mixed species infections are rarely reported in wild-caught mosquitoes. This raises the questions whether simultaneous transmission of both species to mosquitoes is inefficient and whether co-infection may affect ther production of gametocytes or their infectivity. Interestingly, *P. malariae* co-infection is associated with increased *P. falciparum* gametocyte carriage [13] and vice-versa [14]. Whether *P. falciparum* and *P. vivax* interact in ways that modulate gametocyte development, competitive dynamics, or transmission potential remains unknown. In general, the transmission of non-falciparum parasites remains relatively understudied [15].

In this study, we examine whether mixed-species infections alter transmission potential by investigating asexual parasite and gametocyte densities among naturally acquired mixed *P. falciparum–P. vivax* infections and mono-infections and directly assessing transmission to *Anopheles* mosquitoes using mosquito feeding assays.

## Methods

### Ethical approval

Ethical approval was obtained from the AHRI/ALERT Ethics Review Committee (Ref. No. P042/18, and Ref. No. PO-058-24), the National Research Ethics Review Committee (Ref. No. MoSHE/RD/141/1097/19), and London School of Hygiene and Tropical Medicine (LSHTM) (Ref. No. 22518). Before sample collection, all participants and/or the parents/guardians of the children provided written informed consent.

### Study area and patients

This study was conducted between 2022 to 2025 at four health centers in Ethiopia: Shele Health Center and Lante Health Centers in Arbaminch (southern Ethiopia), Maksegnit Health Center in Gondar (northwestern Ethiopia), and Mizan Health Center (southwestern Ethiopia) (**Fig 1**). Across these sites, self-presenting patients with signs and symptoms of malaria were screened, and those with microscopy-confirmed *P. vivax (Pv)* mono-infection, *P. falciparum (Pf)* mono-infection, or mixed*-Pv-Pf* infection were enrolled. Participants received treatment according to national guidelines. Patients infected by *P .vivax* were treated with chloroquine (total dose: of 25mg base/kg for 3 days) plus 14-day primaquine (0·25mg/kg body weight per day). For patients with *P. falciparum* mono-infection or mixed *Pv-Pf* infection, artemether-lumefantrine combined with a single low dose of primaquine was administered.

**Fig 1.**
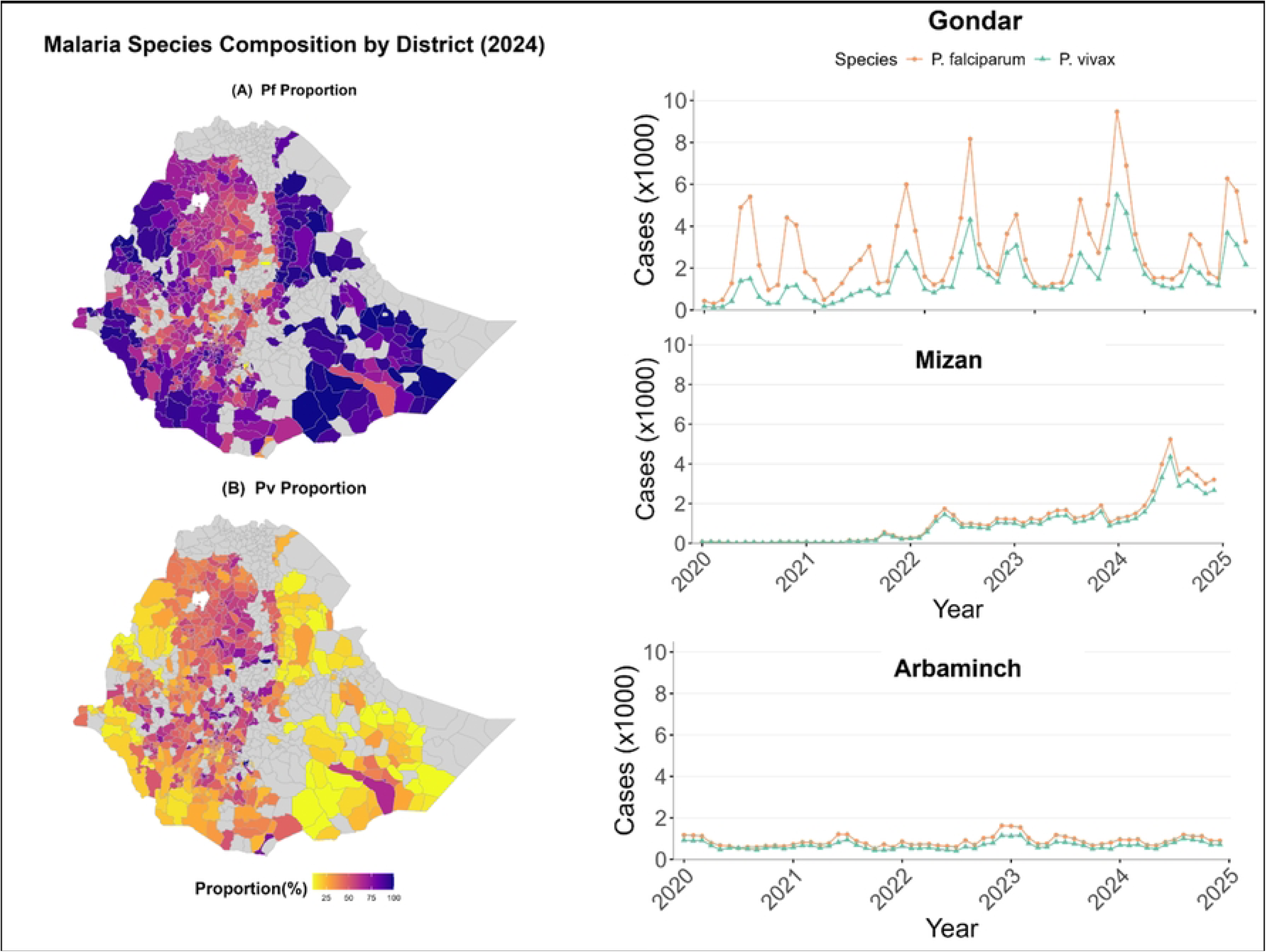
The map of the study sites and malaria case distribution over the past five years (2020 to 2025). In (**A**) shows the overall distribution of malaria cases throughout the country and the contribution of *P. falciaprum* to the overall infection detected per district (proportions of *P. falciparum*, determined by *P. falciparum* cases divided by total malaria cases reported in the district) using data from the 2020 to 2025 Public Health Emergency Management (PHEM) system. Districts with no data available and malaria free are shown in white while more proportion of *P. vivax* infections indicated in color gradient. Shown in (**B**) shows the overall distribution of malaria cases throughout the country and the contribution of *P. vivax* to the overall infection detected per district (proportions of *P. vivax*, determined by *P. vivax* cases divided by total malaria cases reported in the district). Trend line shows the case flow over the past five years for the three selected sites.

### Blood Sample collection and parasite quantification

Prior to treatment, venous blood samples were collected into vacutainer EDTA tubes (using Precision-Glide™ Multi-sample Needles) for parasite and gametocyte quantification and Lithium heparin tubes for Direct Membrane Feeding Assay (DMFA). Blood samples in EDTA tubes were used to extract genomic DNA using Kingfisher Flex robotic extractor (Thermo Fisher Scientific™). DNA was extracted from 50μL whole blood using MagMAX™ magnetic bead-based technology DNA multi-sample kit following the manufacturer’s protocol. Multiplex quantitative PCR (qPCR) targeting the 18S rRNA small subunit gene for *P. falciparum* and *P. vivax* was run using primer and probe sequences described before [16, 17] using TaqMan Fast Advanced Master Mix (Applied Biosystems). *P. falciparum* parasites were quantified using standard curves generated from serial dilutions of NF54 ring stage parasites (10^6– 10^3 parasites/mL). For *P. vivax*, parasite quantification was done using recombinant plasmid constructs to infer copy numbers by running serial dilutions (10^7 – 10^3 copies/mL). Blood samples in protective buffer (RNA Protect Cell Reagent; Qiagen) were used to extract total RNA using the same MagMAX™ magnetic bead technology on a Kingfisher Flex robotic extractor (Thermo Fisher™). Reverse transcriptase quantitative PCR (RT-qPCR) targeting *P. vivax Pvs25* transcript and *P. falciparum* male (*PfMGET*) and female (*CCp4*) gametocyte mRNA transcripts were done [16] using Luna Universal Probe One-Step RT-qPCR Kit (New England Biolabs, NEB). Gametocyte quantification for both species was achieved using *in vitro* RNA constructs in serial dilutions (10^8 – 10^3 copies/mL). Four negative controls were included in each extraction batch in a 96 deep well format for both DNA and RNA extractions. Serial dilutions of the standard curves were generated in duplicate on each plate.

### Direct Membrane Feeding Assay

Blood samples collected in Lithium Heparin tubes were used to feed colony maintained *Anopheles arabiensis* mosquitoes in DMFA following established procedures [18, 19]. All materials used in DMFA were kept at 37°C before sample collection and feeding to mosquitoes. Briefly, blood in pre-warmed heparinized tubes was fed to 3-5 days old female mosquitoes that were starved for ∼12 hours using mini-glass feeders (each 0.3mL capacity) that were covered with PARAFILM® membrane (SIGMA-ALDRICH) and connected to a water bath at 39°C (Laan, Heiloo) for 25 minutes in the dark. Fully blood fed mosquitoes were maintained at 26-28°C and a relative humidity of 60-80% with 10% sugar solution, and were dissected for oocyst detection in the midgut that was stained in 1% mercurochrome on day 7 post feeding for *P. vivax,* and day 10 post feeding on *P. falciparum* and mixed species infected blood fed mosquitoes [20]. Approximately 30 mosquitoes were thus dissected for oocyst detection; additional mosquitoes were kept alive until day 12 post-infection for further molecular analysis of sporozoites and processed if oocyst reading of d7-10 mosquitoes from the same experiment indicated successful transmission. For this sporozoite analysis on day 12 mosquitoes, DNA was extracted following mosquito homogenization by mini-bead beater based as previously described [21]. Briefly, whole mosquitoes were homogenized in 150 µl molecular-grade water with 0.2 g zirconium beads (1 mm diameter) and homogenized using a Mini-Bead Beater-96 (BioSpec). Part of the homogenate (50µL) was used for nucleic acid extraction using cetyl trimethyl ammonium bromide. Then, genomic DNA extracted from the whole mosquitoes was tested on a PCR that targeted 18S small ribosomal subunit gene for parasite detection and quantification following the procedures explained above for blood samples.

### Data Analysis

Statistical analyses were performed using STATA (version 17.0, Stata Corp., TX, USA) and R (version 4.5.1). Proportions were compared using one sample proportion test, and Pearson chi square test or Fisher exact test for independent observations. Equality tests with categorical variables were tested by two-sample Wilcoxon rank-sum (Mann-Whitney) test. Differences among independent groups were tested by Kruskal–Wallis test. Spearman rank correlation coefficient (ρ) was used to assess associations between continuous variables. Continuous variables were presented as medians and interquartile ranges (IQRs). The probability of mosquito infection was modeled as a function of gametocyte density using generalized additive mixed models (GAMMs) with a binomial outcome [22]. The model included log_10_-transformed gametocyte density and infection type (mono vs mixed) as fixed effects, and a random intercept for individual ID to account for repeated measures.

## Results

### Characteristics of study participants

A total of 511 individuals were enrolled between 2022 and 2025 in Arbaminch, Gondar, and Mizan. Of these, 61.8% (316/511) were male, with a median age of 18 years (IQR: 12–25), and 50.9% (220/511) were febrile at recruitment. Microscopy showed substantial misclassification of mixed-species infections. Nearly half (48%; 37/77) of the mixed infections that were detected by PCR, were microscopically classified as mono infections. Consequently, PCR was used for the final classification and resulted in 284 (55.6%) of infections being *P. vivax* mono-infections, 150 (29.4%) *P. falciparum* mono-infections, and 77 (15.0%) mixed *Pv-Pf* infections. No significant differences in age or sex were observed between individuals with mono- and mixed-species infections (**Table 1**).

**Table 1.** Characteristics of patients enrolled in the study.

| Characteristics | *P. vivax* mono infection (N=284) | *P. falciparum* mono infection (N=150) | *P. vivax* and *P. falciparum* mixed (N=77) |
| --- | --- | --- | --- |
| Sex (male); % (n/N) | 60.2<br>(171/284) | 65.3<br>(98/150) | 61.0<br>(47/77) |
| Age (median; IQR) | 15 (11 - 25) | 19 (14-28) | 17 (12-23) |
| Under 5 years, % (n/N) | 0.7<br>(2/284) | 3.3<br>(5/150) | 2.6<br>(2/77) |
| 5 to 15 years, % (n/N) | 44.0<br>(125/284) | 24.7<br>(37/150) | 36.4<br>(28/77) |
| 15 years and above | 55.3<br>(157/284) | 72.0<br>(108/150) | 61.0<br>(47/77) |
| Microscopy agreement with PCR, % (n/N) * | 96.5<br>(274/284) | 98.7<br>(148/150) | 52.0<br>(40/77) |
| Fever at recruitment (axillary temperature $\geq 37.5^{\circ}\text{C}$), % (n/N) | 52.1<br>(148/284) | 50.7<br>(38/75) | 46.6<br>(34/73) |

Abbreviations: IQR: Interquartile Range; NA: Not applicable; \* The number of diagnoses accurately made by microscopy

### Gametocyte densities are higher in *P. falciparum* mono-infections compared to mixed-species infections

Median *P. vivax* parasite density was 54,870 Pv18S copies/µL (Interquartile Range (IQR) 16,131 – 192,977) in *P. vivax* mono-infections and lower in mixed-species infections (median 24,522 Pv18S copies/µL; IQR 9,267 – 66,291; *p*<0.0002; **Fig 2A**). In contrast, *P. falciparum* parasite density did not differ significantly between mono infections (median 7,470 parasites/µL; IQR 1,589 – 33,307) and mixed-species infections (median 5,640 parasites/µL; IQR 755 – 33,356; *p*=0.36; **Fig 2C**). *P. vivax* gametocyte positivity was high and comparable between *P. vivax* mono-infections (97.2%; 276/284) and mixed-species infections (93.5%; 72/77). However, for *P. falciparum*, gametocyte positivity was significantly higher in mono-infections (91.8%; 135/147) than in mixed-species infections (64.5%; 49/76) (*p*=0.001) **(Table 1**). Similar to asexual parasite density, gametocyte density was higher *P. vivax* mono-infections (median 24,160 Pvs25 transcripts/µL; IQR 4,199–115,428) compared to mixed-species infections (median 11,110 Pvs25 transcripts/µL; IQR 2,682–137,242) although this difference was not statistically significant (*p*=0.13; **Fig 2B**). Gametocyte density was markedly higher in *P. falciparum* mono-infections (median, 140 gametocytes/µL; IQR 16–691) compared to mixed-species infections (median 4 gametocytes/µL; IQR 0–107; *p*<0.001; **Fig 2D**). Parasite and gametocyte densities correlated positively in both *P. vivax* mono-infections (ρ=0.40; *p*<0.001) and *P. falciparum* mono-infections (ρ=0.34; *p*<0.001), but these associations were not apparent in mixed-species infections**, (Fig 3A and 3B)**.

**Fig 2.**
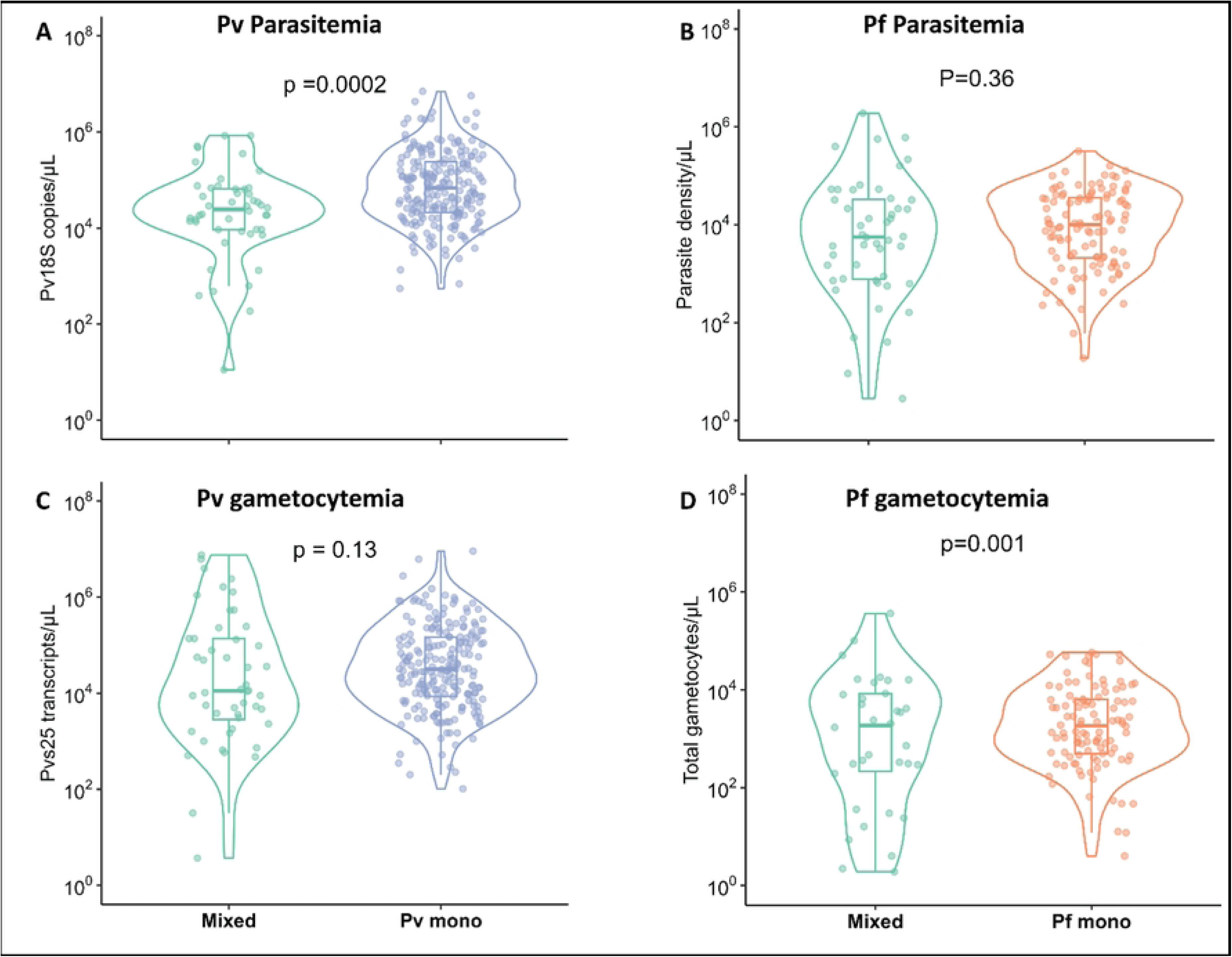
Parasite and gametocyte density comparison and association in the patients’ blood between mono and mixed infections for *P. vivax* and *P. falciaparum*; **A)** Violin plot showing the median parasiste density between *P. vivax* mixed (n=48) and mono infections (284); **B)** Violin plot showing the median gametcoyte density between *P. vivax* mixed (n=48) and mono infections (n=276); **C)** Violin plot showing the median parasiste density between *P. falciparum* mixed (n=48) and mono-infections (n=150); **D)** Violin plot showing the median gametocytemia between *P. falciparum* mixed (n=48) and mono-infections (n=135).

**Fig 3.**
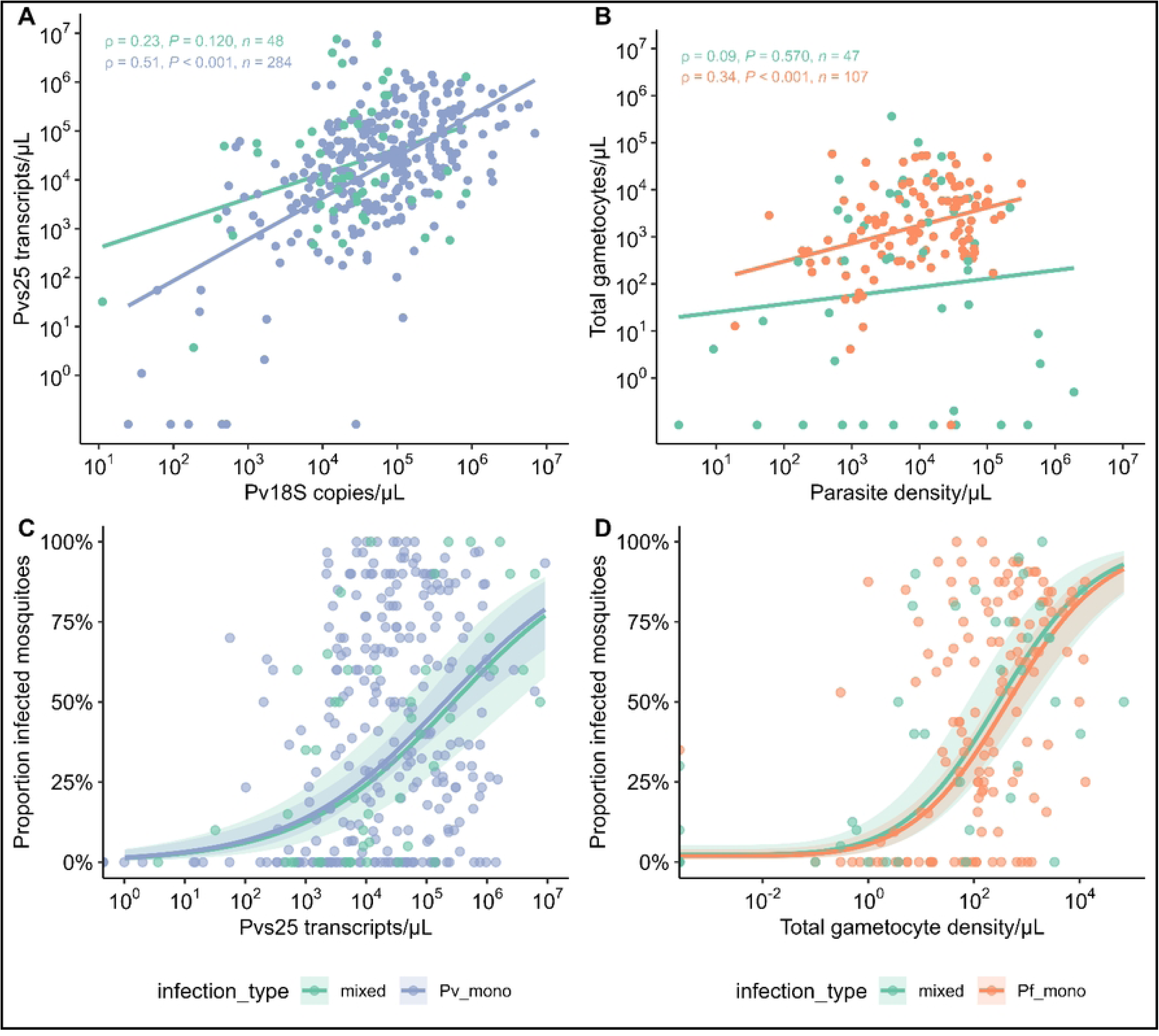
The Generalized Additive Mixed Models (GAMMs) for the prediction of mosquito’s infectivity from gametocytemia in both *P. vivax and P. falciparum* mixed- and mono-infections. **A**) Association between parasitemia and gametocytemia between *P. vivax* mono- and mixed-infections. The x-axis represents log₁₀ Pv18S copies/µL, and the y-axis represents Pvs25 transcripts/µL; **B)** Association between parasitemia and gametocytemia between *P. falciparum* mono- and mixed-infections. The x-axis represents log₁₀ parasites/µL, and the y-axis represents total gametocytes/µL, calculated as the sum of male and female gametocytes/µL; **C)** Proportion of infected mosquitoes (y-axis) against *P. vivax* gametocytemia (Log_10_ transformed Pvs25 transcripts/µL on x-axis); **D**) Proportion of infected mosquitoes (represented on y-axis) against *P. falciparum* total gametocytes/µL, calculated as the sum of male and female gametocytes represented on x-axis. For each infection, the number of infected mosquitoes out of the total dissected was used as the response variable. The model included log_10_-transformed gametocytemia and infection type (mono vs mixed infection) as fixed effects. A random intercept for individual infections was included to account for repeated measures.

### High infectivity to mosquitoes from *Plasmodium* mono- and mixed-species infections

Overall, 74.6% (381/511) of patients were infectious to mosquitoes (i.e. infecting ≥1 mosquito with ≥1 oocyst). The prevalence of infectious feeds in *P. vivax* mono infections was 75.4% (214/284); in *P. falciparum* mono-infections this was 72.0% (108/150) and in *Pv-Pf* mixed infections it was 76.6% (59/77). Across all experiments, 40.8% (6,078/14,884) of mosquitoes became infected with a mean oocyst density of 13 (IQR: 5-37) in infected mosquitoes. The proportion of infected mosquitoes was not different between infection types *P. vivax* mono-infections (39%; 3192/8181) *vs P. falciparum* mono-infections (43.7%; 1978/4522) *vs Pv-Pf* mixed infections (41.6%; 908/2181). Similarly, mean oocyst density was not different (*p*=0.5949); *P. vivax* mono infections (median: 11; IQR: 5 - 32) *vs P. falciparum* mono infections (median: 15; IQR: 5 - 54) *vs Pv-Pf* mixed infections (median: 15; IQR: 4- 53); **Table 2**.

**Table 2.** Mosquito infectivity across infection types and their gametocyte prevalences and desities at oocysts stage.

|  | *P. vivax* infected patients | *P. falciparum* infected patients | *P. vivax* and *P. falciparum* infected patients |
| --- | --- | --- | --- |
| *P. falciparum* gametocyte prevalence, % (n/N) | NA | 91.8<br>(135/147) | 64.5<br>(49/76) |
| *P. falciparum*, gametocytes/μL | NA | 140 (16–691) | 4 (0–107) |
| *P. vivax* gametocyte prevalence, % (n/N) | 97.2<br>(276/284) | NA | 93.5<br>(72/77) |
| *P. vivax*, Pvs25 transcripts/μL (median, IQR); | 24,160 (4,199–115,428) | NA | 11,110 (2,682–137,242) |
| Infectious feeds % (n/N) | 75.4<br>(214/284) | 72.0<br>(108/150) | 76.6<br>(59/77) |
| Infected mosquitoes, % (n/N) | 39<br>(3192/8181) | 43.7<br>(1978/4522) | 41.6<br>(908/2181) |
| Mean oocyst density, median (IQR) | 11<br>(5- 32) | 15<br>(5 - 54) | 15<br>(4 - 53) |

NA: Not Applicable.

To further assess species-specific infectivity in mixed-species infections, 750 mosquitoes from 48 DMFAs using blood samples co-infected with mixed *Pv-Pf* infections were processed on day 12 for sporozoites detection, followed by 18S qPCR analysis of salivary glands. All of these 48 DMFAs resulted in mosquito infections at the oocyst level, as detected in mosquitoes from the same cage that were dissected on earlier days. Among these, 85.4% (41/48) of patients transmitted parasites to at least one mosquito. The majority (65.9%; 27/41) transmitted both species, either concurrently to the same mosquito or to different mosquitoes, while the remainder transmitted only *P. vivax* (21.9%, 9/41) or only *P. falciparum* (12.2%, 5/41). The overall proportion of sporozoite-positive mosquitoes was 51.2% (384/750). Infection rates were highest in mosquitoes carrying both parasite species (67.6%, 261/386) or only *P. falciparum* (67.8%, 61/90), compared to those infected with *P. vivax* only (46.2%, 62/134) (*P*<0.001). *P. vivax* gametocytes were detected in all mixed-species infections (100%; 48/48). Individuals with mixed species infections who transmitted only *P. falciparum* had markedly lower *P. vivax* gametocytemia (median: 1,595 Pvs25 transcripts/µL; IQR: 473–5,327) compared to those who transmitted only *P. vivax* (median: 48,974 Pvs25 transcripts/µL; IQR: 15,000–96,807; *p*<0.0001) and those who transmitted both species (median: 11,958 Pvs25 transcripts/µL; IQR: 3,600–1,100,000; *p*<0.001). *P. falciparum* gametocyte positivity (11.1%; 1/9) and density (70 gametocytes/µL in a single donor) were very low in those individuals who transmitted *P. vivax* only, while it was comparable between those only transmitting *P. falciparum* (median; 257 gametocytes/µL; IQR: 83–465) and those transmitting both species (median; 407 gametocytes/µL; IQR:7–1,765; *p*= 0.8493). Overall, non-infectious individuals exhibited lower gametocyte densities for both species (**Table 3; S1 Fig)**.

**Table 3.**
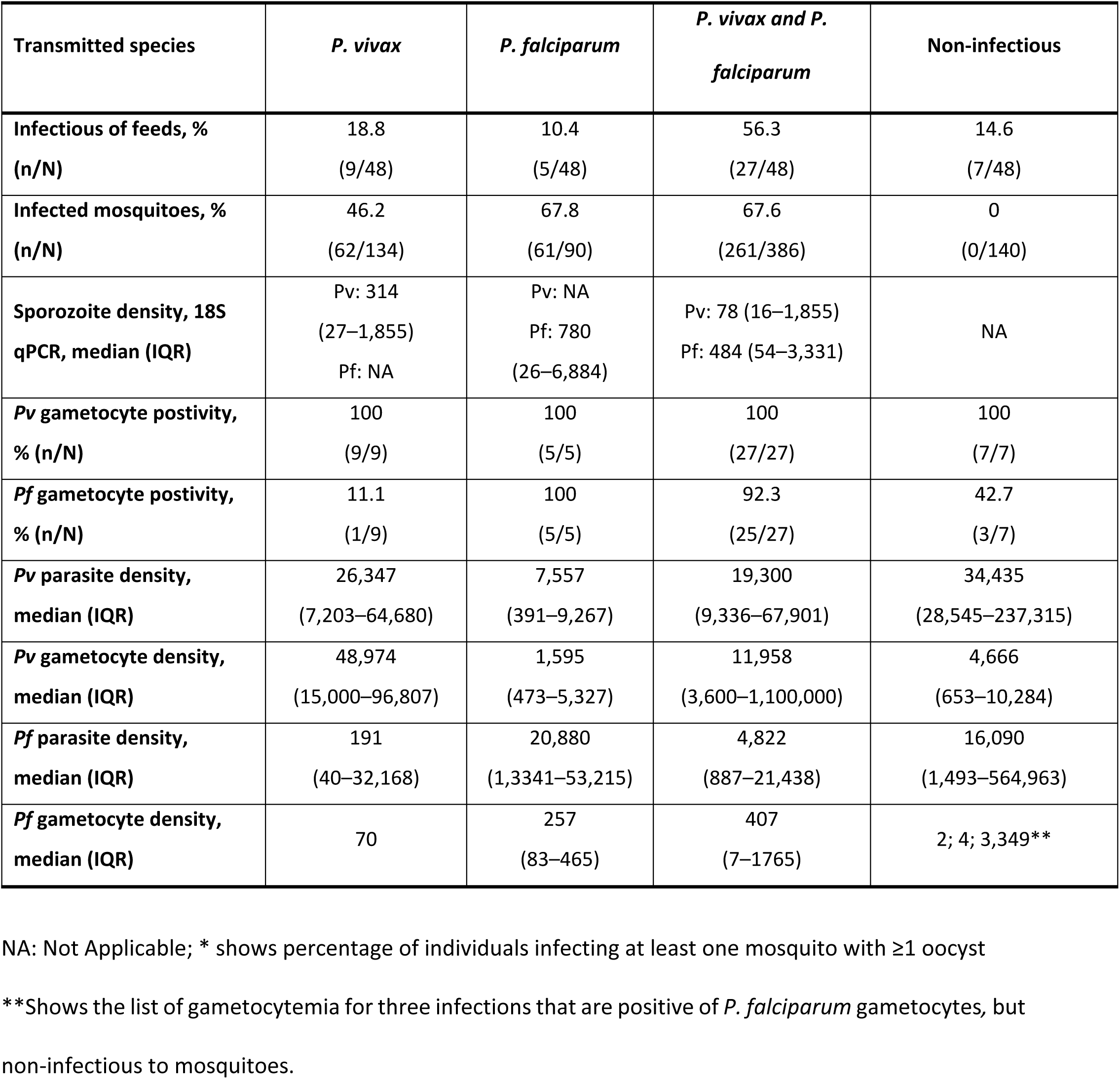
Characteristics of mixed-species feeds with sporozoite outcomes.

Finally, among individuals who transmitted both *P. vivax* and *P. falciparum,* most had high gametocyte densities for both species, facilitating dual transmission. Notably, 96.3% (26/27) of these patients with dual transmission, transmitted both parasites to single mosquitoes, although some mosquitoes carried only one of the two species. Sporozoite density, as determined by 18S qPCR, did not differ between those who infected with *P. vivax* mono- and mixed-species infections. Similarly, sporozoite density was comparable between mosquitoes infected with *P. falciparum* mono-infections and mixed-species infections (**S2 Fig).**

### Gametocyte density is the most important determinant of transmissibility to mosquitoes

The probability of mosquito infection was positively associated with gametocytemia for both *P. falciparum* and *P. vivax*, in both mono- and mixed-species infections. Using generalized additive mixed models (GAMMs), we predicted mosquito infection rates based on gametocyte density. For *P. falciparum*, each log₁₀ increase in gametocytemia was associated with a nearly threefold increase in the odds of mosquito infection (OR=2.96; 95% CI: 2.36–3.71; *p*<0.001), with no significant difference observed between mono-and mixed-species infections (OR=0.82; 95% CI: 0.40–1.69; *p*=0.596). Similarly, *P. vivax* gametocytemia was positively associated with proportion of infected mosquito (OR=2.26; 95% CI: 1.74–2.93; *p*<0.001; for each log increase), again with no significant difference between mono- and mixed-infections (OR=1.11; 95% CI: 0.58–2.15; *p*=0.774) (**Fig 3C and 3D; S1 Table)**.

## Discussion

This study provides direct evidence on the relative infectiousness to mosquitoes of natural mixed species *Plasmodium* infections in co-endemic settings. *P. falciparum* gametocyte positivity and density were lower in mixed infections; *P. vivax* gametocyte prevalence was uniformly high but density appeared lower in the presence of *P. falciparum*. Most importantly, we observed no evidence for an effect of co-infection on mosquito infection rates after adjustment for gametocyte densities; transmission success strongly correlated with gametocytemia without an impact of a second infecting *Plasmodium* species.

Parasite densities differ markedly between *P. falciparum* and *P. vivax* infections. *P. vivax* parasitemia is generally low due to its strict preferences for reticulocytes that represent only 1–2% of circulating erythrocytes [23, 24]. This limited niche makes *P. vivax* highly vulnerable to factors that reduce reticulocyte availability. *P. falciparum* might exacerbate this constraint through its association with anemia [25] and dyserythropoiesis [26], potentially restricting reticulocyte availability. By comparison, *P. falciparum* has a competitive advantage due to its broader invasion range (young and mature erythrocytes) and higher multiplication rate (16–32 merozoites per schizont *vs* 12–24 for *P. vivax*) [27, 28]. Together, these mechanisms may explain why expecially in mixed-species infections, *P. vivax* parasite density is low.

Despite differences in parasite density, *P. vivax* gametocyte prevalence was similarly high in mixed and mono-infections, indicating efficient gametocyte production regardless of parasitemia [29]. Gametocyte density broadly followed asexual parasite densities and was lower in mixed-species infections. In contrast, *P. falciparum* showed lower gametocyte positivity and density in mixed infections. Whilst it is complicated to draw conclusions on the underlying mechanism in the absence of longitudinal data, this finding appears consistent with prior observations that co-infection may suppress *P. falciparum* gametocyte carriage [14, 30].

Uniquely, we did not only measure parasite densities in human blood but also directly assessed the transmissibility of infections to mosquitoes. For both species, mosquito infectivity increased sharply with gametocyte density [31]. The ‘per gametocyte infectivity’ was not altered by the presence of an additional, and potentially competing, parasite species. Whether an individual with a mixed-species infection infected mosquitoes with only one or with both *Plasmodium* species was generally well-explained by species-specific gametocyte densities.

We regularly observed that both parasites were transmitted, sometimes to the same mosquito. This suggests that mosquitoes are highly receptive to both species, even if present in the same bloodmeal, and suggest that there may be no competition within the mosquito vector, a finding supported by previous work showing concurrent transmission from mixed-species infected blood samples [32]. The finding that we observed no indications for parasite competition within mosquitoes suggests that this is not an (important) explanation for the lower prevalence of *P. vivax* infected mosquitoes that is commonly observed in field collections [33] or the lower incidence of *P. vivax* infections in populations living in co-endemic settings. The higher proportion of vectors carrying *P. falciparum* alone or with *P.vivax* could be (partially) explained by blood-stage interactions. *P. falciparum*-induced anemia and dyserythropoiesis – both not examined in this study - can reduce reticulocyte availability, limit *P. vivax* expansion [34]. Immune-mediated effects and resource competition may further disadvantage *P. vivax,* even when gametocytogenesis is preserved [35, 36].

Although we report that dual transmission occurs, including both parasites being detected within single mosquitoes, mixed-infections remain rare in wild-caught *Anopheles* [37, 38]. This scarcity likely reflects multiple factors: asynchronous gametocyte production between species, diagnostic under-detection of minority infections [11]. If *P. vivax* infections would typically be of shorter duration than *P. falciparum* infections, this could explain why mosquitoes are rarely detected with both sporozoite species.

However, the sparse studies that examined infection duration in co-endemic settings suggest that *P. vivax* infections may in fact last longer than *P. falciparum* [39].

Our analysis relied on naturally acquired infections and mosquito feeding assays that provide biologically relevant estimates of human-to-mosquito infectivity. As a limitation, our findings were based on single time-point observations of parasite density, gametocyte density and mosquito infection rates. Our experiments thereby do not answer all questions on species interactions. We observed differences in parasite densities in co-infections but exert caution in the interpretation of these differences since we did not prospectively monitor asexual parasite and gametocyte kinetics and therefore cannot conclude whether co-infection alters parasite multiplication rates, gametocyte conversion rates or infection longevity. These questions would require a different (longitudinal) study design and may be examined in future studies. We conclude that there is no evidence for competition within mosquito hosts with co-infection not altering the association between gametocyte density and mosquito infection rates for either Plasmodium species.

## Acknowledgments

We thank the study participants who volunteered to participate in the study. We also thank the study team members at field sites and malaria program officers at the regional, zonal, and district health offices who supported the execution of the study.

## Authors’ Contributions

Conceived and designed the study protocol: FGT, TB, WC;

Carried out the sample collection: DN, AG, WC, KG, KA, AJK, BG, ZS, TB, TE, MD;

Carried out the molecular experiments (DNA extraction qPCR and RT-qPCR): WC, TA, GJ, GH, MA, AS, MKB;

Carried out data analysis and interpretation of results: LAE, SS, WC, FAK, MD;

Drafted and reviewed the manuscript: WC, FGT, LAE, HM, FM, SC,TB, CD;

All authors critically revised the manuscript for intellectual content, read and approved the final manuscript.

## Data and code availability

All the data used in the paper will be made available on Dryad (linked with the ORCID: https://orcid.org/0000-0003-1931-1442). Raw data from the study will be available in the future upon written request to the corresponding author following the signing of the data-sharing agreement, abiding by institutional and international data-sharing guidelines. The R codes used to run the analyses reported in this study will be made available at (GitHub link).

## Supporting information Captions

**S1 Fig. *P. falciparum* and *P. vivax* gametocyte densities for mixed species feeds.** *P. vivax* (blue) and *P. falciparum* (orange) gametocyte densities for mixed *P. vivax* and *P. falciparum* infections offered to mosquitoes in DMFAs are indicated in box plots. The whisker plot indicates the distance between the Interquartile range values in the dataset with the middle line indicating the median value of the gametocyte density.

**S2 Fig. Comparison of the parasite density in mosquitoes at sporozoite stage**. It was determined by 18sqPCR between mono and mixed species infected mosquitoes for *P. vivax* and *P. falciparum*.

**S1 Table.** The Odds Ratio of mosquitoes’ infection rate based on gametocyte density for both *P. vivax* and *P. falciparum* mixed infection and mono infections.

| Variabls | OR (95% CI) | *p*-value |
| --- | --- | --- |
| *P. falciparum* infectivity |  |  |
| Gametocyte density increase (log<sub>10</sub> scale) | 2.96 (2.36–3.71) | <0.001 |
| Reference (*P. falciparum* mixed infection) |  |  |
| *P. falciparum* mono infections | 0.82 (0.40–1.69) | 0.596 |
| *P. vivax* infectivity |  |  |
| Gametocyte density increase (PvS25 copies/μL) (log<sub>10</sub> scale) | 2.26 (1.74–2.93) | <0.001 |
| Reference (*P. vivax* mixed infections) |  |  |
| *P. vivax* mono infections | 1.11 (0.58–2.15) | 0.774 |

